# Registration and medical certification of deaths in the Indian States: A comparative analysis of data of CRS and MCCD reports (2010-2019)

**DOI:** 10.1101/2021.12.09.21267291

**Authors:** Anuj Kumar Pandey, Diksha Gautam, Benson Thomas M, Yogita Kharakwal

## Abstract

**Background:** The medical certification of cause of death (MCCD) under Civil Registration System (CRS) has been implemented in the States/UTs in a phased manner to provide data on cause of death but due to incomplete coverage and inadequate quality of civil registration data and medically certified data system, use of this data has been compromised. The completeness of registration of death (CoRD) and completeness of medically certified deaths were assessed from 2010 to 2019 at state level to understand their current status and trend over time and also to identify gaps in data to improve data quality.

**Methods:** CoRD and CoMeRD for each year for each state was calculated from the CRS reports and MCCD reports respectively for the period 2010-2019. Data were analyzed nationally as per geographical region and individual state. Union Territories excluding Delhi and Telangana have not been considered in this analysis.

**Results:** The CoRD in India have increased in the CRS from 66.9% in 2010 to 92 percent in 2019, a significant increase of 37.7% over 9 years (P<0.001) whereas India has not witnessed a substantial increase in the CoMeRD in MCCD which has increased from 17.1% in 2010 to only 20.6% in 2019. Among the 29 States, 18 (62%) had CoRD >95 percent in 2019, with 15 states recording 100 percent of CoRD however just 3 states (10.3%) have CoMeRD more than 50% namely Goa (100%), Manipur (67.3%) and Delhi (61.7%).

**Interpretation & conclusions:** Despite the significant progress made in CoRD in India, importance of medical certification cannot be undermined; critical differences between the States within the CRS and MCCD remain a cause of concern. Concentrated efforts to assess the strengths and weaknesses at the State level of the MCCD and CRS processes, quality of data and plausibility of information generated are needed in India.

## Background

The demand for better health statistics is growing at a very rapid pace, including both empirical data and estimates on various aspects of health such as births, deaths, morbidity, risk factors, health systems, and health service coverage. These parameters are the basis for evidence-based decision-making with regard to resource allocation, monitoring of indicators, identifying the priorities for programs and other related activities in the area of public health (1,2).

The Civil Registration System (CRS) in India is generating these vital statistics(3). The Medically Certified Cause of Death (MCCD) reports are the series of the publication presenting statistics on causes of death obtained through the Civil Registration System under the Registration of Births and Deaths Act, 1969(2). In many developing countries including India, the civil registration data are not following the data quality parameters of completeness and also not available in time and therefore, compromising the usefulness of these data. (4)

The non-availability of reliable and quality data of mortality, morbidity and cause specific deaths are a major concern as they result in countries incapable of tracking and safeguarding the well-being of their populations due to unavailability of the records of millions of deaths and births in the country(5).

The Government of India under Section 10(2) and 10(3) of the Act empowers the State Government to enforce the provision relating to medical certification of cause of death in specified areas taking into consideration the availability of medical facilities and this act also empowers the States to issue a certificate of the cause of death by the medical practitioner who has attended to the deceased at the time of death(2).

In view of the registration of birth and death in the CRS, India implemented registration of a sample of births and deaths as part of the Sample Registration System (SRS) in 1970(6) which is the main source of vital statistics including cause of death for India. The Sample Registration System (SRS) provides reliable annual estimates of Infant Mortality Rate, birth rate, death rate and other fertility & mortality indicators at the national and subnational levels. It is a large-scale demographic survey conducted every year by the Office of the Registrar General, India in all States and Union Territories(7)

The MCCD under Civil Registration System has been implemented in the States/UTs in a phased manner to provide data on cause of death. However, its implementation is not uniform in all the States/UTs. It is still being implemented mostly in urban areas as selected by the Chief Registrar of Births & Deaths. So, the scheme covers mostly those deaths occurring in medical institutions located in urban areas. The coverage under the scheme in terms of percentage level of medical certification as well as the type of hospitals covered has not been uniform across the States/UTs(2).

The Registrar General India (RGI) has been impressing upon all the States/UTs for increasing the registration of deaths and despite a notable increase of 41% in medically certified registered death in India in the past 3 decades with the increase in reporting by States/UTs but ??the States/UTs?? are still struggling with the quality of the available data (Fig: I).

**Fig I:**
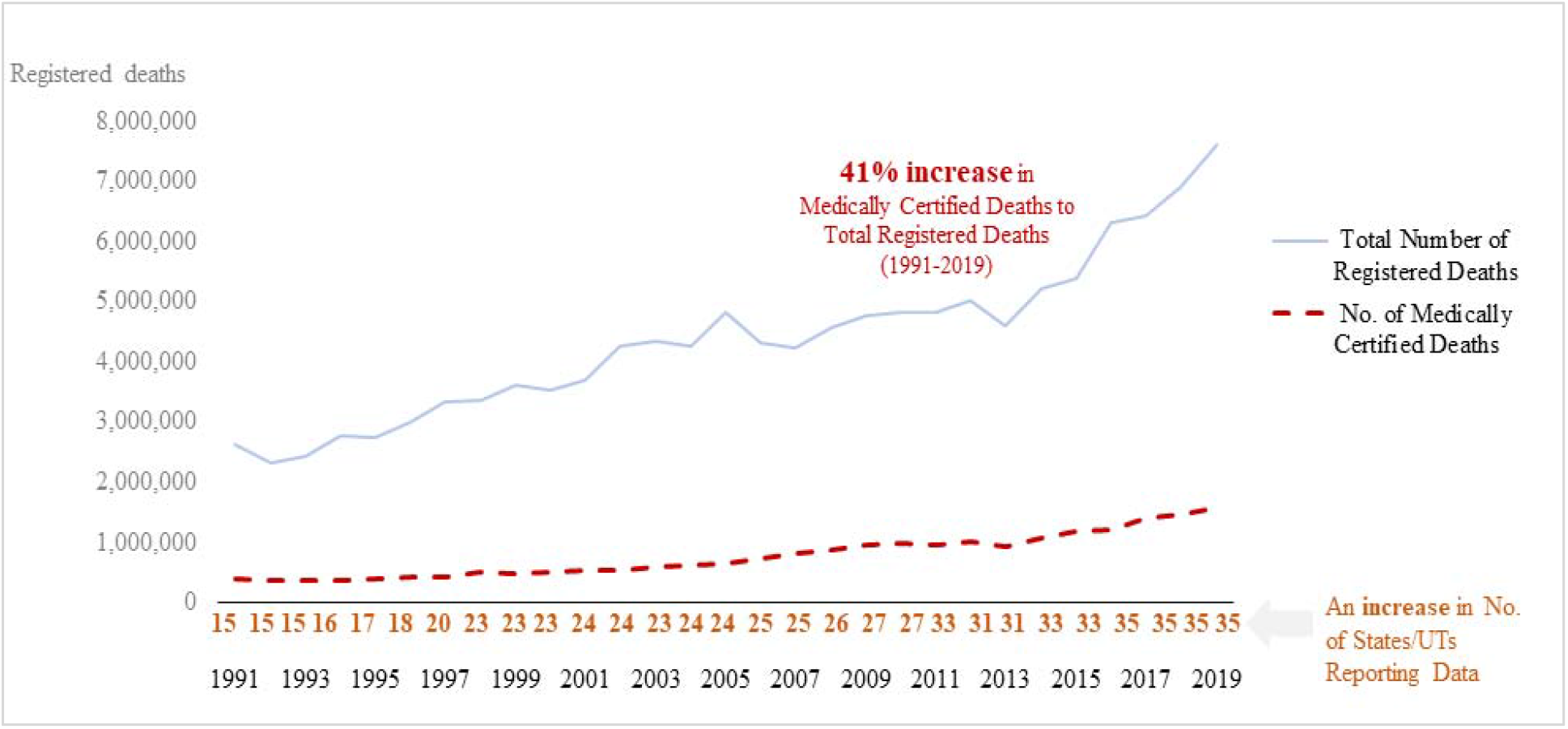
Showing growth in Medical Certification as a part of total registered deaths; Source: MCCD report 2019(2)

So, the study explored completeness of death registration in the CRS and MCCD report in India from 2009 (or is it 2010?) to 2019 with the aim to get an insight on the trends and current status of completeness of death and medically certified death registration.

The aim is to assess death registration nationally as per geographic region and as per individual States with the aim of highlighting the inconsistency in death registration that could facilitate prioritization of immediate actions to improve coverage, and to identify the gaps in data to improve CRS data quality. The secondary objective is to look for the status of completeness of medically certified deaths in Indian States. It is important to mention that the CRS in India does not regularly document information on the cause of death which is supplemented by SRS.

## Material and Methods

The data related to medically certified deaths registration have been taken from Medical Certification of Cause of Death (MCCD) report which is a fundamental part of the Vital Statistics System. This provides a reliable and sequential database for generating cause-specific mortality statistics(2).

For this study, the ‘Vital Statistics of India based on Civil Registration System’ was used which provided information on the completeness of death registration for each State for a given year(3). The administrative reporting system of a birth/death event in India starts at the local level; the consolidated registrations from the local level are transmitted to the chief registrar of a State from where an annual consolidation of these data is sent to the Office of the Registrar General and Census Commissioner of India (RGI), Government of India. Based on this State-level annual consolidation, the RGI office produces the CRS reports annually for each State(2,3).

These reports which are publicly available were downloaded each year from 2009 to 2019 as the most recent year available at the time of this analysis was 2019. The death and medically certified deaths registration details were compiled from these reports for analysis. The data on estimated deaths were also fetched from CRS report that is available on its website.

The available data were analyzed to find out the longitudinal trends in completeness of registration of deaths (CoRD) and Completeness of Medically certified Registered Deaths (CoMeRD) for the years from 2009 to 2019 from the CRS and MCCD reports respectively, and the change in coverage from 2009 to 2019 for India overall, as per geographical regions, and as per each State were reported.

The CoRD in the CRS report is defined as the percentage of registered deaths to the deaths estimated through SRS for a given year(3) whereas we have also calculated CoMeRD which is defined as percentage of medically certified deaths to total registered deaths in MCCD report(2). India was categorized into six geographical regions based on the SRS classification(6). The State wise progress from 2009 to 2015 and then 2019 were reported. The geographical region coverage was calculated as the average coverage of all States in that region.

All analyses were carried out using MS Excel 365. The z-test was applied where relevant to assess significance in univariate analysis. The union territories along with Telangana were excluded, Delhi was considered as a State.

## Result

The CoRD in India have increased in the CRS from 66.9 percent in 2010 to 92 percent in 2019, a significant increase of 37.7 percent over 9 years (P<0.001) whereas India has not witnessed a substantial increase in the CoMeRD in MCCD which has increased from 17.1% in 2010 to 20.6% in 2019. (Fig-II)

**Fig II:**
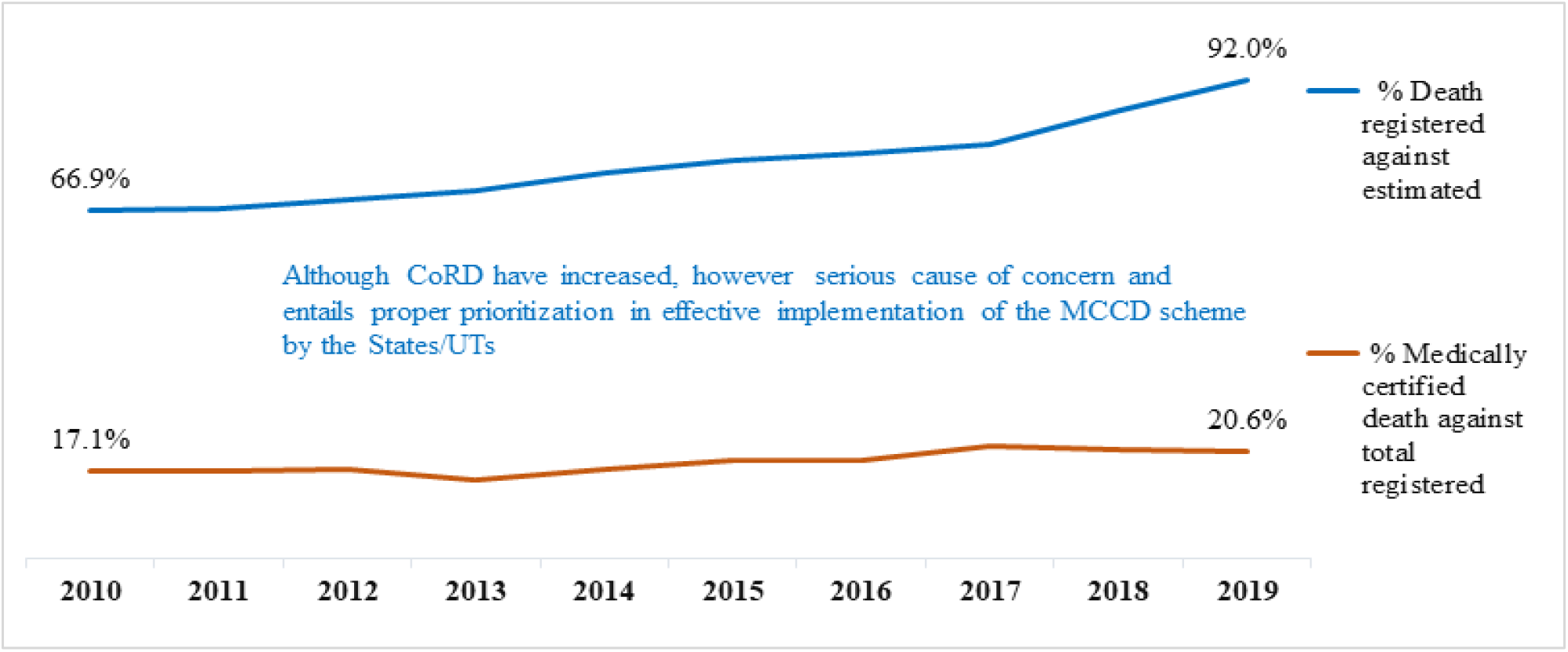
Status of Completeness of Registration of Death (CoRD) vis-a-vis completeness of medically certified registered deaths (CoMeRD) from 2009 to 2019(2,3)

**Fig III:**
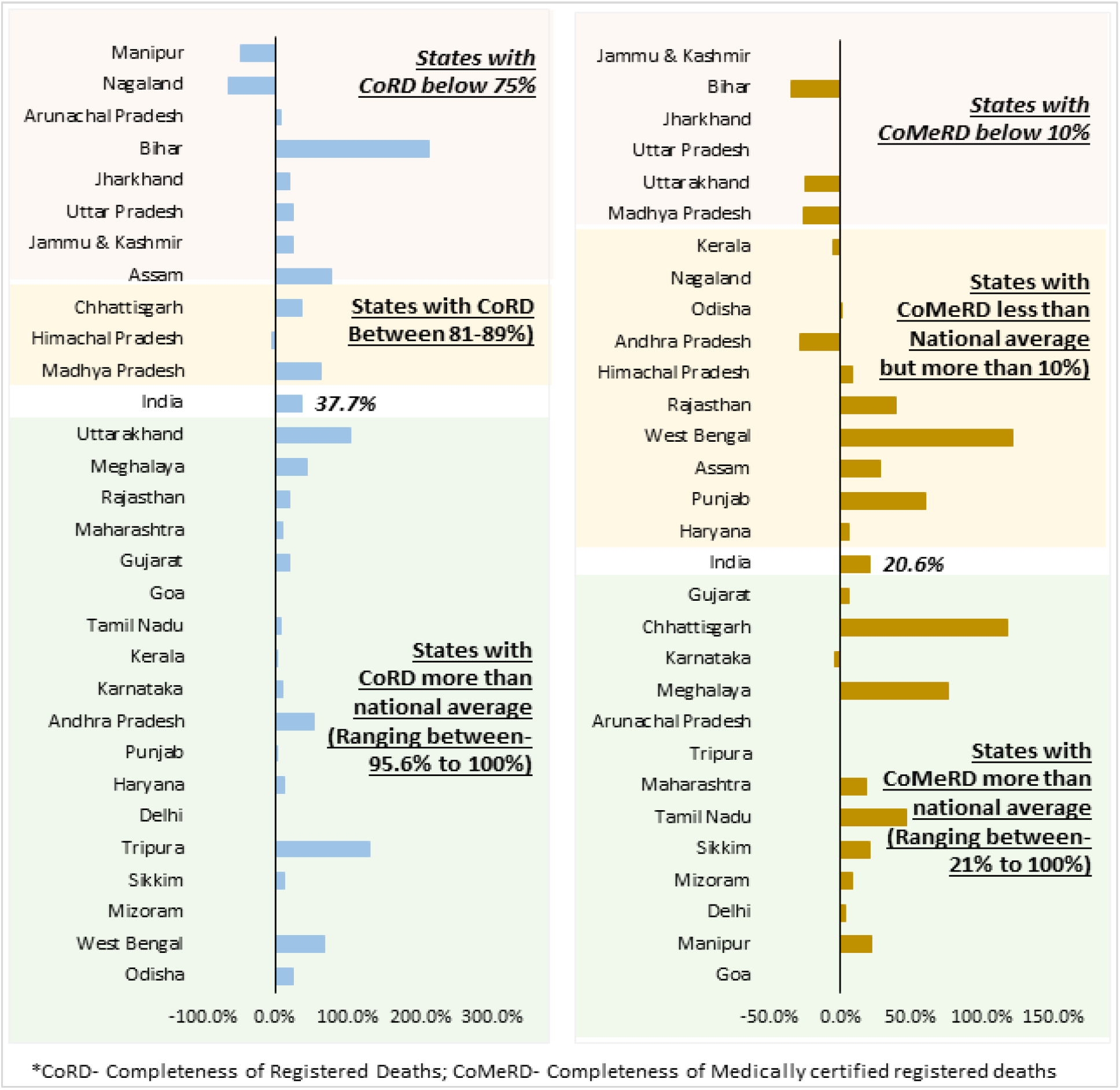
Percentage change in completeness of Registration of Death (CoRD) and completeness of medically certified registered deaths (CoMeRD) from 2009 to 2019(2,3)

There is a heterogeneous increase in CoRD across most States during 2010 to 2019. Considering the geographic regions, in 2019; Western region has achieved 100% of registration of deaths against estimated deaths and the same region have also reported more than 50% (∼53.2%) of medically certified registration in deaths against total registered deaths.

The CoRD is >90% (National average ∼92%) in North, South and Western geographic regions leaving central (83.1%), Eastern (77.6%) and Northeastern (70.6%) regions behind. Highest percent change in the geographic regions was documented in the eastern region (50.8%) and change in the northeastern region still needs special attention (11.6%) as shown in (Table I)

**Table I:** Completeness of Registration of Deaths (CoRD) in 2010, 2015 and 2019 and per cent change from 2009 to 2019 for India, the geographic regions, and each Indian State in the Civil Registration System.

| *Geographic Region/States\** | *CoRD* |  |  | *Per cent change in CoRD between 2010 and 2019* |
| --- | --- | --- | --- | --- |
|  | *2010 (in %)* | *2015 (in %)* | *2019 (in %)* |  |
| **India** | **66.9** | **76.6** | **92.0** | **37.7** |
| **Central** | **62.0** | **68.8** | **83.1** | **34.2** |
| *Chhattisgarh* | 60.1 | 87.3 | 81.5 | 35.7 |
| *Madhya Pradesh* | 54.8 | 53.8 | 89.1 | 62.5 |
| *Rajasthan* | 82.4 | 89.9 | 98.6 | 19.7 |
| *Uttar Pradesh* | 50.6 | 44.2 | 63.3 | 25.1 |
| **East** | **51.5** | **68.9** | **77.6** | **50.8** |
| *Bihar* | 16.5 | 31.9 | 51.6 | 212.5 |
| *Jharkhand* | 49.4 | 70.1 | 58.8 | 19.0 |
| *Odisha* | 80.0 | 100.0 | 100.0 | 25.0 |
| *West Bengal* | 59.9 | 73.5 | 100.0 | 67.0 |
| **North-East** | **62.9** | **61.3** | **70.2** | **11.6** |
| *Arunachal Pradesh* | 35.7 | 46.6 | 38.6 | 8.1 |
| *Assam* | 41.8 | 51.1 | 74.0 | 77.0 |
| *Manipur* | 40.7 | 34.0 | 21.4 | -47.6 |
| *Meghalaya* | 67.5 | 78.2 | 97.6 | 44.7 |
| *Mizoram* | 100.0 | 100.0 | 100.0 | 0.0 |
| *Nagaland* | 85.1 | 29.2 | 30.0 | -64.7 |
| *Sikkim* | 89.1 | 100.0 | 100.0 | 12.2 |
| *Tripura* | 43.5 | 51.4 | 100.0 | 129.7 |
| **North** | **79.3** | **85.8** | **91.5** | **15.4** |
| *Delhi* | 100.0 | 100.0 | 100.0 | - |
| *Haryana* | 88.6 | 100.0 | 100.0 | 12.9 |
| *Himachal Pradesh* | 89.7 | 89.0 | 86.4 | -3.7 |
| *Jammu & Kashmir* | 53.4 | 60.4 | 66.7 | 24.9 |
| *Punjab* | 97.0 | 100.0 | 100.0 | 3.1 |
| *Uttarakhand* | 46.9 | 65.4 | 95.6 | 103.7 |
| **South** | **86.9** | **92.5** | **99.4** | **14.4** |
| *Andhra Pradesh* | 64.8 | 84.9 | 100.0 | 54.2 |
| *Karnataka* | 91.1 | 96.2 | 100.0 | 9.8 |
| *Kerala* | 99.2 | 100.0 | 100.0 | 0.8 |
| *Tamil Nadu* | 92.6 | 100.0 | 100.0 | 8.0 |
| **West** | **91.3** | **99.2** | **100.0** | **9.5** |
| *Goa* | 100.0 | 100.0 | 100.0 | - |
| *Gujarat* | 82.6 | 100.0 | 100.0 | 21.1 |
| *Maharashtra* | 91.4 | 97.5 | 100.0 | 9.4 |

\*Chandigarh, D & N Haveli, Daman & Diu, Lakshadweep, Puducherry, A & N Islands and Telangana removed from analysis.

The medically certified deaths against total registered deaths (CoMeRD) have not increased substantially over these years but Northeastern region has reported the highest (∼67%) change in CoMeRD. North, South, and Western regions need a rigorous follow-up to increase the coverage of this scheme. (Table II)

**Table II:**
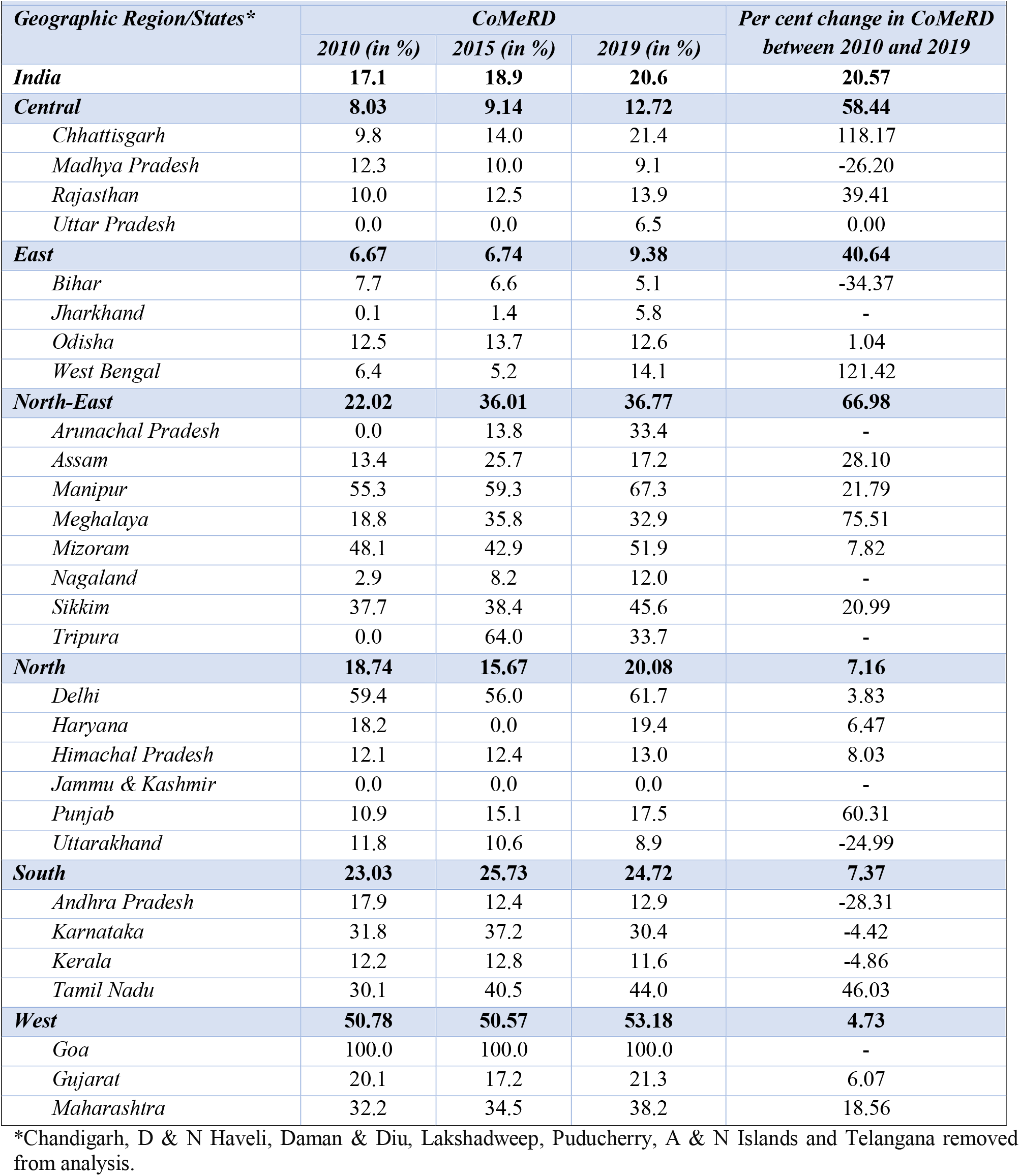
Completeness of Medically certified Death Registration (CoMeRD) in 2010, 2015 and 2019 and per cent change from 2009 to 2019 for India, the geographic regions and each Indian State in the MCCD.

Among the 29 States, 18 (62%) had CoRD >95 percent in 2019, with 15 States recording 100 percent of CoRD (Table-I) however just 3 states (10.3%) have CoMeRD more than 50% namely Goa (100%), Manipur (67.3%) and Delhi (61.7%). Except Mizoram, Sikkim and Tripura other north North-eastern States have the lowest CoRD as compared to the national average (92%) with Manipur (−47.6%) and Nagaland (−64.7%) showing a decrease in CoRD between 2009 to 2019. Comparing the CoMeRD, except Nagaland (12%) and Assam (17.2%) all the other north-eastern States have higher CoMeRD as compared to the National average (∼20.6%) showing a better acceptance and reporting of medical certification of deaths against total registered deaths. (Table: I & II)

Highest percent increase (212.5%) has been noticed in Bihar with CoRD of 51.6% percent. Except for some North-Eastern States, changes in Bihar (51.6%), Jharkhand (58.8%), Uttar Pradesh (63.3%), and Jammu & Kashmir (66.7%) remained almost static and are still some of the States of concern where the death reporting against estimated is lower as compared to other States. (Table: I)

As per the data, on one side 100% deaths in Goa are medically certified whereas some of the states namely Jammu and Kashmir (0%), Bihar (5.1%), Jharkhand (5.8%), Uttar Pradesh (6.5%), Uttarakhand (8.9%), Madhya Pradesh (9.1%) have <10% medical certification. Lastly 9 states (31%) namely Bihar (−34.4%), Andhra Pradesh (−28.3%), Madhya Pradesh (−26.2%), Uttarakhand (−24.9%), Kerala (−4.9%), Karnataka (−4.4%) and Uttar Pradesh (0%) have shown a decrease in CoMeRD between 2009 to 2019 (Table: II)

## Discussion

Despite the existence of a robust system for collecting the death registration data, a lot of inconsistencies are present in the completeness of death registration and medically certified cause of deaths. The study was conducted to assess the death registrations and their medical certification in the country, by geographic region and at State level. The data emphasized on the trends in progress made in death registration and medically certified cause of deaths and their heterogeneity at State level in India over a decade. The findings of this could be further utilized to understand the gaps and foster the immediate action plan to improve the CRS data quality as well as the quality of MCCD data(8).

The completeness of death registration has always been a challenge in India but now it is evident from the findings of the study that we have achieved a good milestone of registering more than 90% of deaths at national level which shows the progress in the strict monitoring and review mechanism of deaths. But the concern remains for medical certification as less than one-fourth (20.6%) of deaths in India are getting medically certified.

Among the 29 States, 18 (62%) had CoRD >95 percent in 2019, with 15 States recording 100 percent of CoRD however just 3 states (10.3%) have CoMeRD more than 50% namely Goa (100%), Manipur (67.3%) and Delhi (61.7%).

The continuously low CoRD in the economically weaker States like Bihar (51.6%), Jharkhand (58.8%), Uttar Pradesh (63.3%), and Jammu & Kashmir (66.7%) as well as States which have shown the substantial decline in CoRD like Nagaland and Manipur are alarming and there is an urgent need for improvement in the death reporting system in these States. In the same way, on one side 100% deaths in Goa are medically certified whereas some of the states namely Jammu and Kashmir (0%), Bihar (5.1%), Jharkhand (5.8%), Uttar Pradesh (6.5%), Uttarakhand (8.9%), Madhya Pradesh (9.1%) have <10% medical certification.

There is a big gap of complete and timely death registration in India, which hides the actual extent of the mortality in some states(9). All the states are different from each other in terms of their geography, demography, and economy, therefore, the action to be taken for improvement of death registration must be done at State level rather than at central level(10,11). Lessons from the states which were lagging behind and made progress could be taken to implement in the poorly performing States(9)

On the one hand, while completeness in death registrations is showing definite improvements, on the other hand, the reporting of medically certified deaths shows a major weakness in Indian reporting??? system. The study showed no significant change in medically certified death registered against total registered deaths. There is need to strengthen the system at the level of data collection as even after more than 3 decades of implementation of the program, the coverage is not even one-fourth (∼20.7%) of the total registered deaths. In order to take the scheme of MCCD forward in a systematic manner in the country, the Registrar General, India has to impress upon all the States/UTs to bring all hospitals (whether public or private) and private medical practitioners under its coverage, both in rural as well as urban areas(2,4).

Also, even the deaths which are medically certified and reported, the quality of the certification and assessing the real cause of death is highly compromised and ill-defined, depriving the data of analyzing the real situation of the causes of mortality. Imprecise cause of death in medical certification has also been observed in some of the Medical College hospitals of India(4,12). The cause of death should be reviewed in the presence of an expert physician thoroughly along with monthly review meetings.

In India, a lot of factors acts as the barriers in the death registration and MCCD such as termination of the pension system post death of a retired public sector employee etc.(4). Poor system of initial notification of deaths, deprived mechanism of institutional death registration at village level, inadequate and inefficient monitoring mechanism, absence of digital death registration mechanism, inadequate training and sensitization workshops at community level, insufficient and incompetent human resource, poorly functioning health facilities and registration units are some other factors adding to poor death registration in rural states(13).

This study recommends the following steps to improve the death registrations and medically certified death reporting in India. There is need of creating awareness in the communities on the benefits of death registration, strengthening of the system of initial notification of deaths, implementing strict and better monitoring of death registration data, strengthening capacity building of staff, and making registration offices more readily accessible(13). Other measures to improve registration includes digitalization of the data entry of deaths and flow of cause of death at both Municipality and State levels, strengthening of the private sector reporting on deaths and MCCD, and joint review meeting of Municipalities and health departments.

## Conclusion

The present analysis concludes that although the death registration over the year have increased significantly, it is required to be given sufficient and long overdue priority to improving the medical certification of the registered deaths. Improvement in the death registration will reduce its dependence on SRS for birth and death estimations. The importance of the medical certification of deaths cannot be undermined for various evidence-based decision-making and for the benefit of public health policy making. Intense effort in the assessment of strengths and weaknesses at the State level of CRS as well as MCCD process needs to be taken to understand and overcome the difficulties at implementation level.

## Data Availability

This study involves only openly available human data, which can be obtained from: https://censusindia.gov.in/2011-Common/mccd.html and https://censusindia.gov.in/2011-Common/mccd.html

## Reference

1. Shibuya K, Scheele S, Boerma T. Health statistics: time to get serious. Bull World Health Organ. 2005 Oct;83(10):722.

2. Census of India WebsiteJ: Office of the Registrar General & Census Commissioner, India [Internet]. [cited 2021 Nov 26]. Available from: https://censusindia.gov.in/2011-Common/mccd.html

3. Civil Registration System [Internet]. [cited 2021 Nov 26]. Available from: https://crsorgi.gov.in/annual-report.html

4. Kumar GA, Dandona L, Dandona R. Completeness of death registration in the Civil Registration System, India (2005 to 2015). Indian J Med Res. 2019 Jun;149(6):740–7.

5. Setel PW, Macfarlane SB, Szreter S, Mikkelsen L, Jha P, Stout S, et al. A scandal of invisibility: making everyone count by counting everyone. The Lancet. 2007 Nov 3;370(9598):1569–77.

6. Census of India: Sample Registration [Internet]. [cited 2021 Nov 26]. Available from: https://censusindia.gov.in/Vital_Statistics/SRS/Sample_Registration_System.aspx

7. SRS Bulletin 2019.pdf [Internet]. [cited 2021 Nov 26]. Available from: https://censusindia.gov.in/vital_statistics/SRS_Bulletins/SRS%20Bulletin%202019.pdf

8. World Health Organization - 2013 - Strengthening civil registration and vital statist.pdf [Internet]. [cited 2021 Nov 30]. Available from: https://www.who.int/healthinfo/CRVS_ResourceKit_2012.pdf

9. Basu JK, Adair T. Have inequalities in completeness of death registration between states in India narrowed during two decades of civil registration system strengthening? Int J Equity Health. 2021 Aug 30;20(1):195.

10. United Nations. Principles and recommendations for a vital statistics system: Revision 3 [Internet]. UN; 2014 [cited 2021 Nov 30]. (Statistical Papers (Ser. M)). Available from: https://www.un-ilibrary.org/content/books/9789210561402

11. Hart JD, Sorchik R, Bo KS, Chowdhury HR, Gamage S, Joshi R, et al. Improving medical certification of cause of death: effective strategies and approaches based on experiences from the Data for Health Initiative. BMC Med. 2020 Mar 9;18(1):74.

12. Pandya H, Bose N, Shah R, Chaudhury N, Phatak A. Educational intervention to improve death certification at a teaching hospital. Natl Med J India. 2009 Dec;22(6):317–9.

13. UNICEF ROSA Status of Civil Registration and Vital Statistics in South Asia Countries 2019.pdf [Internet]. [cited 2021 Nov 30]. Available from: https://www.unicef.org/rosa/media/3121/file/UNICEF%20ROSA%20Status%20of%20Civil%20Registration%20and%20Vital%20Statistics%20in%20South%20Asia%20Countries%202019.pdf

